# Modeling non-pharmaceutical interventions in the COVID-19 pandemic with survey-based simulations

**DOI:** 10.1101/2021.04.16.21255606

**Authors:** Marius Kaffai, Raphael H. Heiberger

## Abstract

Governments around the globe use non-pharmaceutical interventions (NPIs) to curb the spread of coronavirus disease 2019 (COVID-19) cases. Making decisions under uncertainty, they all face the same temporal paradox: estimating the impact of NPIs before they have been implemented. Due to the limited variance of empirical cases, researcher could so far not disentangle effects of individual NPIs or their impact on different demographic groups. In this paper, we utilize large-scale agent-based simulations in combination with Susceptible-Exposed-Infectious-Recovered (SEIR) models to investigate the spread of COVID-19 for some of the most affected federal states in Germany. In contrast to other studies, we sample agents from a representative survey. Including more realistic demographic attributes that influence agents’ behavior yields accurate predictions of COVID-19 transmissions and allows us to investigate counterfactual “what-if” scenarios. Results show that quarantining infected people and exploiting industry-specific home office capacities are the most effective NPIs. Disentangling education-related NPIs reveals that each considered institution (kindergarten, school, university) has rather small effects on its own, yet, that combined openings would result in large increases in COVID-19 cases. Representative survey-characteristics of agents also allow us to estimate NPIs’ effects on different age groups. For instance, re-opening schools would cause comparatively few infections among the risk-group of people older than 60 years.

## 1 Introduction

In 2020, governments tried to curb the spread of COVID-19 without having an appropriate medical response available. Most governments implemented non-pharmaceutical interventions (NPIs) which affect societal life in an unseen manner. Although there is initial evidence that NPIs, such as the closure of educational facilities [7, 26, 27], workplaces [27, 38] and certain businesses [7, 26], the cancellation of mass gatherings [7, 26, 27], case detection and contact tracing [23, 38], shielding of vulnerable people [24], wearing masks [36] or specific network intervention strategies [39] are effective in reducing the amount of new cases, it is also clear that NPIs come with severe side effects, e.g., on the economy [39, 41], labour markets [11], people’s physical [34] and mental health [8], or concerns on increasing domestic violence [6]. Thus, decision-maker act under immense pressure to balance public health risks while sustaining economic activities. In so doing, policy-makers are facing a temporal paradox when it comes to the implementation of NPIs: they have to decide whether a particular NPI (or a set thereof) is curbing the spread of COVID-19 without having reliable knowledge about the NPIs’ actual effectiveness. The best way to evaluate the effectiveness of NPIs would be to measure the empirical correlation between the implementation of a NPI and the change of the infection rate. However, an empirical estimation is difficult in many areas due to the unavailability of data. Only few studies exist with a rather narrow set of scenarios [7, 26, 27] or coarse resolutions [14]. Empirical studies on the effect of NPIs are by-design limited in scope because they can only measure the impact of what governments implement, i.e., estimations rest on scenarios which have actually happened. Based on inter-temporal cross-country data, Brauner et al. [7], for instance, cannot disentangle the effect of school closure and the effect of university closure, because in most countries these two institutions were closed almost at the same time. To circumvent one’s reliance on detailed empirical data in order to evaluate a larger variety of scenarios, simulation methods can be used to create and investigate the effect of NPIs which did not occur in reality [22, 44].

In addition, there is great need for more detailed knowledge of how NPIs work in the specific context of a country’s or region’s social structure [12, 44]. Preliminary evidence exists that the spread of COVID-19 cases is starkly influenced by structural circumstances, such as age distributions [13, 17] or network densities of geographic areas [31]. To reflect social structures and relevant characteristics of people’s behavior in simulations, agent-based modeling (ABM) have proven to be good complements to epidemiological differential equation models [18, 19, 39, 44]. Once a model with ‘predictive capability’ is established [40], it is possible to create scenarios that differ from empirical scenarios but to apply the simulation results to reality [22, 44].

Many researcher already exploit the benefits of data-driven agent-based models to investigate NPIs in the context of a countries’ specific circumstances [9, 10, 20, 24, 30, 35, 38]). ABM exist for the U.S. [10, 20], Canada [38], Australia [9], France [24], Singapore [30], Iran [35] and the UK [20]. Our model not only adds (parts of) Germany to this list but uses a innovative method to generate an artificial population of agents which resembles the real population of a federal state and, hence, important parts of its social structure. Thus instead of corresponding to a few, rather coarse macro-characteristics of the target population, our study represents households of survey participants. The micro-level of the simulation – agents with specific attributes living together with other agents with specific attributes – therefore reflects important properties of the social structure of a society; much more than synthesizing the artificial population based on aggregated macro-data on the target populations’s household structure.

Utilizing large-scale agent-based simulations with representative survey data in combination with SEIR models provides the frame for testing counterfactual “what-if” scenarios in a quasi-experimental setup. This allows us to assess the effectiveness of NPIs for a variety of otherwise unobservable scenarios and deduce the effect of each NPI on its own (cf. “Methods”4 for details). In addition, the micro-foundation of ABMs paves the way to differentiate NPIs’ impact on demographic sub-groups, of which age is the most important in regard to COVID-19 [17, 37].

## 2 Results

To derive the efficacy of NPIs in counterfactual scenarios, the first step is to ensure the predictive capability of the model by assessing its fit to the actual spread of COVID-19. The proposed model matches the empirical data of the daily number of cumulative cases for each of the four most affected federal states in Germany (Figure 1). Our model predicts the rapid rise of COVID-19 cases in the early stages of the pandemic as well as its subsequent decelerated growth in which the infection numbers stabilize. The model also simulates accurately the different levels of disease spread in the four states, which vary considerable by structure (cf. SI Table 3).

**Figure 1:**
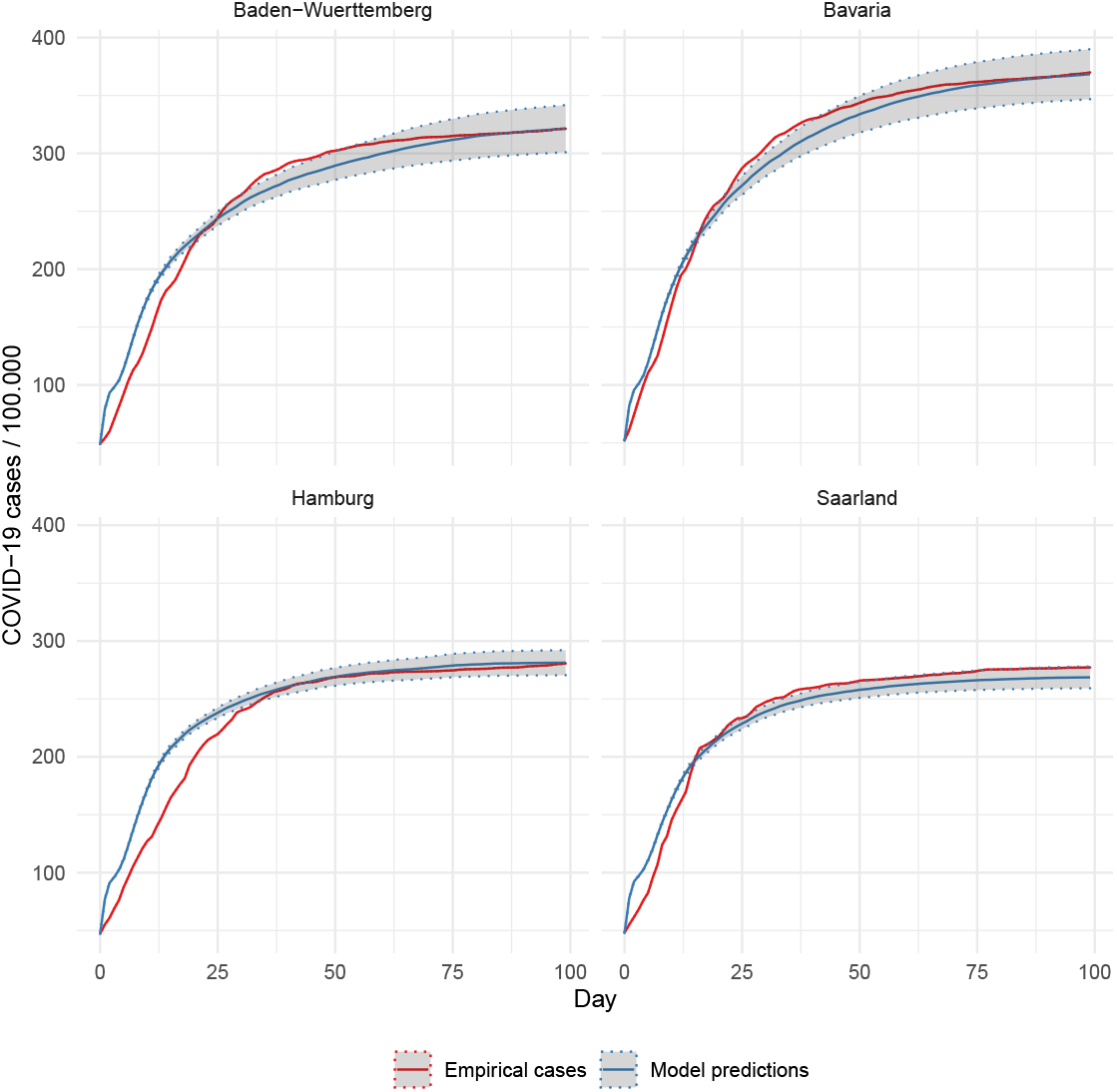
Model fit by state. The solid lines depict the number of reported COVID-19 cases and the average of infections across simulation runs. Shaded areas represent the 95% confidence intervals of the averages. The root mean squared error of the model’s predictions ranges between 10.17 for Bavaria and 17.68 for Hamburg.

After establishing that the model actually predicts the empirical spread of the Corona virus, we can turn to implementing counterfactual scenarios. We do that by removing each NPI that was at work in reality (the “Baseline”-scenario) and measure the effect of its omission in the simulation. Figure 2 reveals that decreasing the urge to quarantine infectious cases and reducing home office hours to regular percentages would cause the starkest increase across all implemented NPIs. While quarantine is, by a margin, the most important NPI in the two large territorial states (Baden-Wuerttemberg and Bavaria), it is a bit less important than increased home-office in the two smaller states. At the other end of the efficacy spectrum is the scenario in which there was no reduction of work hours in certain economic sectors due to economic downturns and the partial shutdown of their business. Across all states, it has almost no effect in our simulation if people would have worked normal hours.

**Figure 2:**
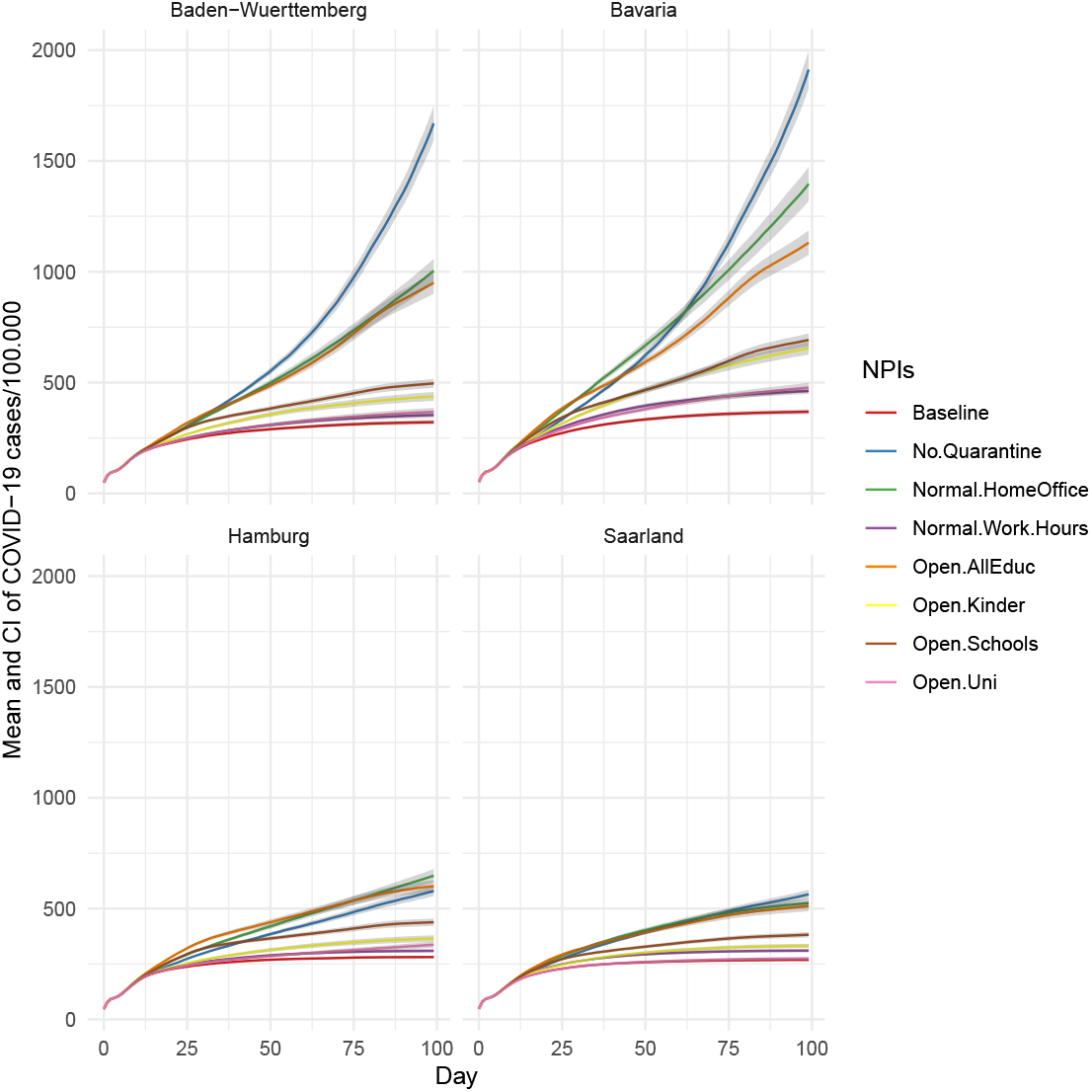
Effect of counterfactual non-implementation of NPIs, by state. Each line represents the effect that the removal of one specific NPI has on the spread of COVID-19 in our simulation. For instance, “Normal.Homeoffice” means that the percentage of people working at home is set to the statistical average, i.e., the value in non-pandemic times.

Which education facilities may be kept running is one of the most controversial topics in Germany, since it affects the lives of millions of pupils and parents. Turning to education-related NPIs, our model predicts a large increase of COVID-19 cases if all three types of institutions (kindergartens, schools, and universities) would re-open. In contrast to previous studies [7], our model also allows us to inspect the effect of each education facility separately. In the counterfactual scenarios presented in Figure 2, opening schools would have increased COVID-19 cases by several hundred (per day). Figure 3 disentangles the numbers further. It shows that opening all education-related institutions at the same time has a considerable effect (third largest overall, 2), however, each institution alone causes much less infections. Therefore, contagious effects seem at work which can be mostly attributed to the interplay of kindergarten kids and pupils (”Open.Schools Kinder” in Figure 3). If comparing education-related NPI individually, we see that the effects of all three types are rather similar. Yet, opening universities has slightly lower effects than the other two, and opening schools causes the highest average effect in all four states.

**Figure 3:**
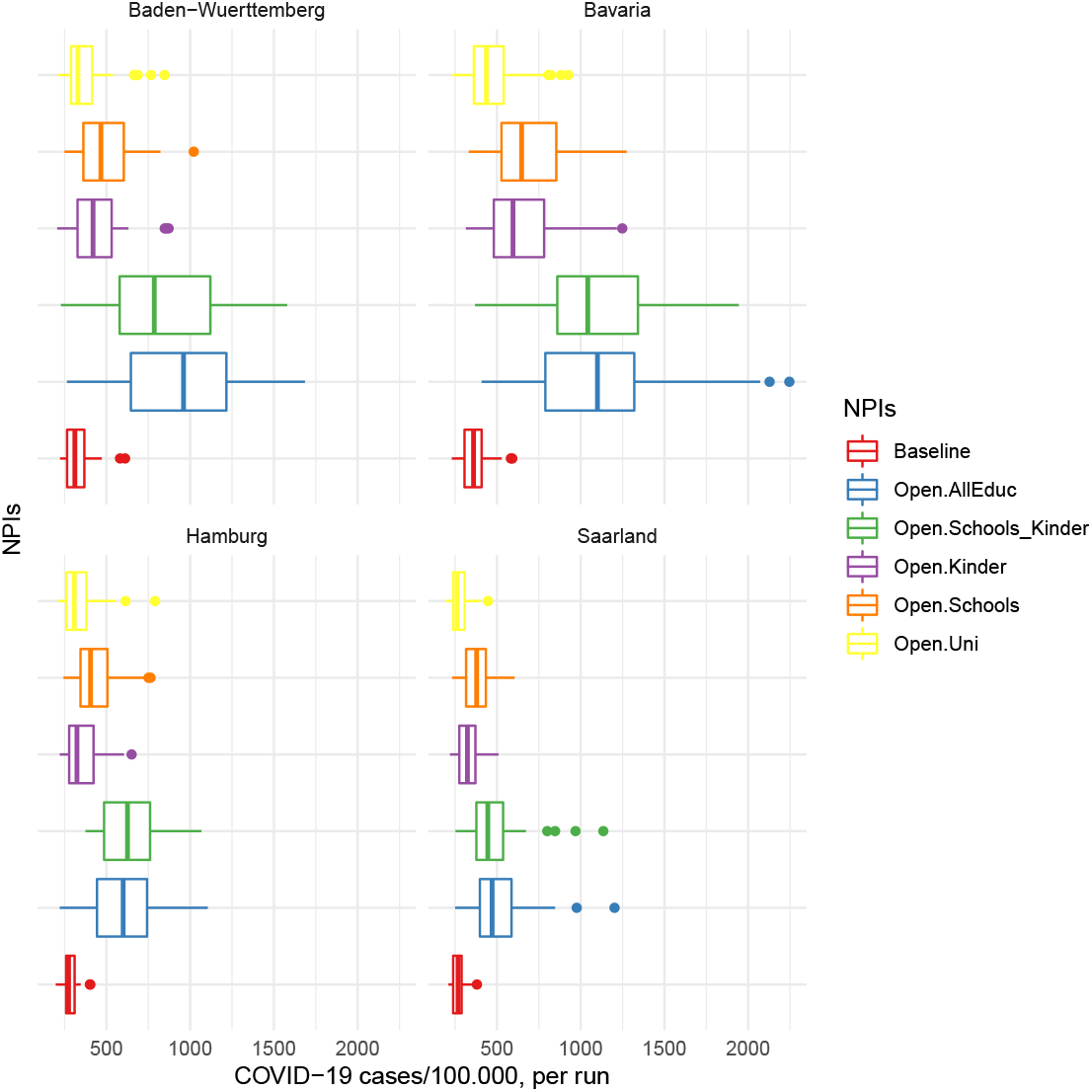
Cumulative infection numbers for each counterfactual non-implementation of education-related NPIs, by state. Boxplots depict the distribution of all runs.

Besides considering NPIs separately, using an ABM with representative survey respondents as agents allows us to trace infections by age group. Age constitutes one of the main factors to identify risk groups [17]. In general and for each NPI, the mean age of infected agents decreases over time (Figure 4). This reflects recent findings using rare empirical data on the age of infected [37]. Furthermore, Figure 4 details how each NPI affects different age groups. It reveals that the counterfactual removal of NPIs drive COVID-19 numbers through the infection of agents younger than 60. Across all states, the scenarios in which schools re-open and pre-pandemic home office rates are attained lead to the lowest proportion among the elderly risk-group. In contrast, re-opening the universities yields a higher proportion of infections among people older than 60, although the vast majority of infections pertains younger people and the absolute numbers of COVID-19 caused by re-opening universities are rather low (Figure 2).

**Figure 4:**
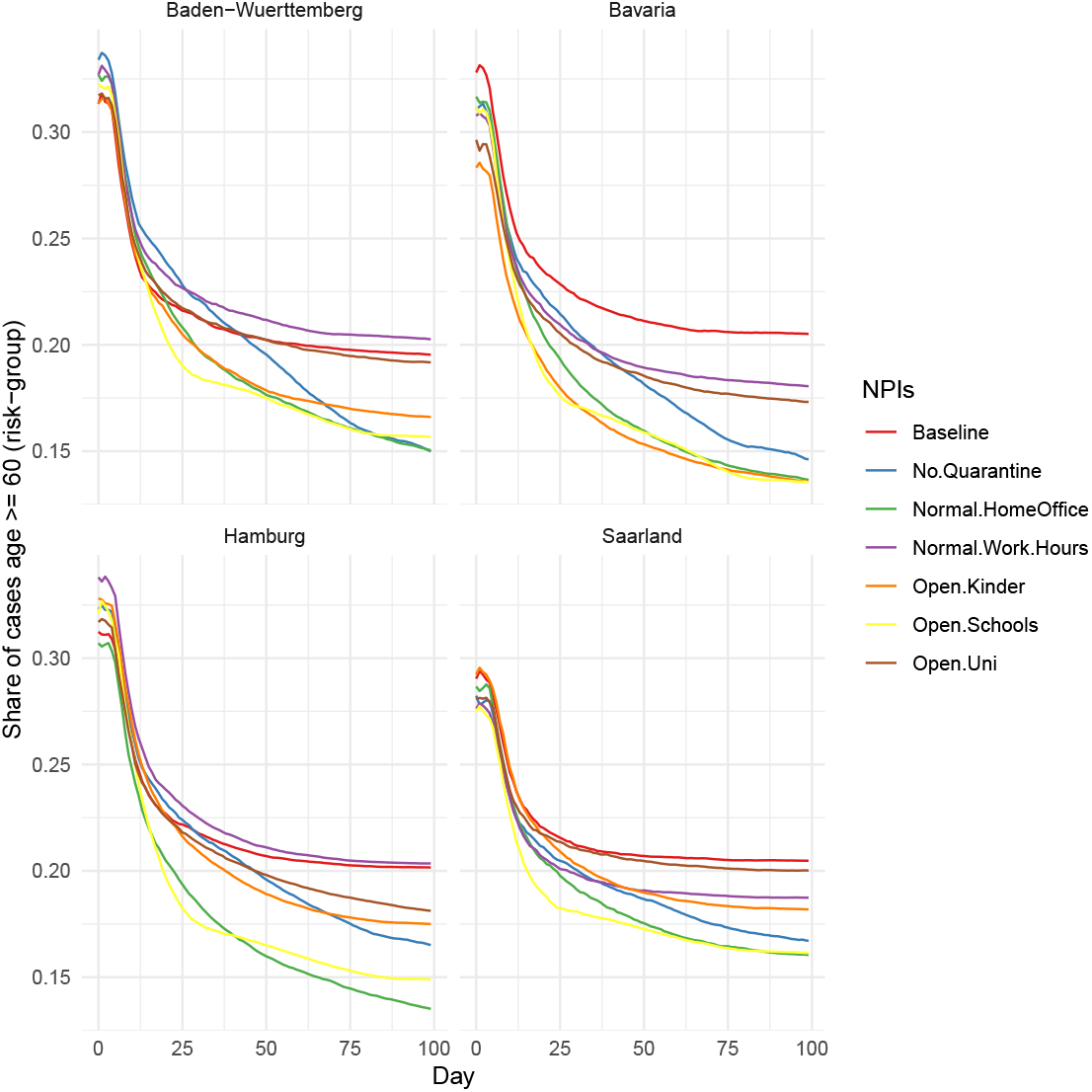
Number of infected agents belonging to the risk-group (age >= 60), by state. Lines represent the share of cases in the risk group for each NPI over all simulation runs. To facilitate readability, a ribbon of confidence intervals is omitted. Figures including the CIs are available upon request.

## 3 Discussion

Overcoming the temporal paradox of deciding whether the implementation of a particular NPI is decreasing the spread of COVID-19 before knowing its actual effect is crucial for policy-makers and researchers alike. Testing such counterfactual “what-if” scenarios could improve the basis for decision-making considerably and help to decide which NPIs to implement. Our approach illustrates a way to estimate the effectiveness of otherwise unobservable scenarios by simulating agents with attributes taken from a large survey representing about 27 million people. In so doing, our study generates insights on the effectiveness of several widespread NPIs (quarantine, home-office, closing universities, schools, and kindergartens, reduced work hours) in four federal states in Germany. Going beyond previous studies, our approach enables us to disentangle the effect of individual NPIs and, hence, provides information for policy makers who need to balance the trade-off underlining each NPI between individual and economic freedom on the one side, and prevention of disease spread on the other.

It can be shown that focused quarantining of infected agents and their household members (i.e., close contacts) is the most effective NPI. Governments should therefore make every effort to increase their ability of case detection and contact tracing [23, 38]. The second largest effect is the home office scenario. It can be shown that an increased usage of home office capacities is a very effective measure to reduce the number of new infections, which is in line with observational studies [3]. This result is particularly important, because although there was a considerable increase of home office usage in the beginning of the pandemic [32], both employers and employees still refuse to comprehensively rely on home office in the long term [29].

The most controversial NPI is the closure of educational facilities. In contrast to many observational studies [7, 14, 21], we are able to estimate a separate effect for each education-related NPI. Our simulations suggest that the separate re-opening of each institution yields rather low infection numbers. However, combined openings lead to multiple infections, in particular, re-opening kindergartens and schools at the same time increase COVID-19 cases considerably. Therefore, alternating periods of open educational institutions could represent a promising way to maintain low infection numbers while sustaining institutional functions.

A key characteristic of COVID-19 is its elevated mortality for older people and the subsequent importance of demographics [17]. Taking into account social structures by using agents with survey-characteristics and in accordance with a very recent study for the USA [37], our findings reveal that younger age (less risk-prone) groups sustain the spread of COVID-19 when the fore-mentioned NPIs are omitted. Furthermore, simulations suggest that the removal of NPIs affect age groups differently. Considering age adds an important puzzle piece how to select NPIs, i.e., it allows to distinguish the impact of NPIs in regard to its overall effects *and* group-specific effects for the people who are most vulnerable to COVID-19.

Like other simulation studies, our analysis has several limitations. First, our model does not contain a representation of contacts between households or agents that do no take place in one of the implemented locations, e.g., contacts with relatives or friends. Second, our model does not contain locations were people meet during spare time. However, the influence of both points is mitigated because private contacts and leisure activities were reduced to a minimum in the observed period.

A further limitation is that we do not model changes of people’s behavior. For example, we do not take into account the change in caution about infection risks in daily life or the obligation to keep distance and wear masks inside buildings. Also beyond the scope of this paper is the inclusion of external influences. Instead, we simulate a closed system. After the initial infections at the beginning of a run no infections occur due to agents coming home from “outside”, e.g., commuters or tourists.

As many other simulation studies, we had to estimate some parameters due to a lack of empirical data. However, we conducted a series of robustness checks for the most important of those parameters (cf. Figures 5-9 in SI). Sensitivity analyses suggest that the results are robust to changes in the parameters and, hence, to potential errors in assumed values.

Despite those limitations, we provide a reproducible model based on empirical agents which can be adopted and/or extended (Python code freely available). For instance, the current model including above NPIs could be readily applied on any states or regions. Researchers would only need to calibrate the model with subsequent empirical data on COVID-19 cases and survey data. In addition, researcher could extend the existing model with alternative NPIs which may be relevant for a specific region. The simulated world can be amplified by many other “meeting places” in which certain persons come into contact.

Despite all efforts and (slowly) increasing availability of vaccinations, it appears that COVID-19 will accompany our societies for some time to come [33]. Thus, the demand for data-driven epidemiological models including realistic social structures and demography will stay high. Our model is an instance how to incorporate survey-based populations and test counterfactual scenarios. The development and improvement of such models and the timely availability of more accurate data was maybe never more important than in the current global pandemic.

## 4 Methods

### Creating the population of agents

Each agent-based model consists of 100000 agents living their live during the pandemic.^1^ They are spending their time either at home, at an occupation specific work place, at kindergarten, at school, at university or at a supermarket. Each action depends on the day of week, the time of day and individual characteristics of each agent. The values of the agents’ characteristics, such as age, gender, occupation, daily working hours or the time usually spent in the supermarket, are taken from survey data, i.e., each agent represents a survey respondent in regard to his or her attributes. For that purpose, we use the 2017 German Socio-Economic Panel (SOEP) [43]. The SOEP is a representative panel survey that includes entire households of respondents. Table 1 (cf. supplementary information SI) lists all variables used to set agents’ attributes.

We model the infection on the level of federal states with differing social structures. We chose the four states in Germany (Baden-Wuerttemberg, Bavaria, Hamburg, Saarland) with the highest infection rate relative to its population size during the first wave of Covid-19 in Germany (March 2020). While including other states/regions with lower infection numbers is possible, in principle, it is difficult to apply on our closed system because the disease spread dies most often in early stages without the influx from outside.

To create a population of agents that represents a given federal state, we first create a subset of SOEP-respondents who reside in the given state. In a second step, we take a random sample (with replacement) from the subset until a population size of at least 100000 is reached. The households’ selection probabilities are not equal, but are weighted by the corresponding cross-sectional representativity weight provided by the SOEP for each household.

Like every survey, SOEP has to deal with missing values. For 3.53% of the sample we replace missing data on working hours by age- and gender-specific group means. For 3.46% of the sample we replace missing data on the occupation by a random occupation code in each simulation run. For cases with missing information about the time spent daily in supermarkets (26,92%) we impute the overall mean (approximately 1h). Finally, 1.62% of the households are excluded, because the case itself or a member of the corresponding household has missing data.

To set the agents’ daily routines in the simulation, we classify each agent either as kindergartner, school student, working, university student, working university student or non-working agent. Therefore, we use “empirical agents” to trace the spread of COVID-19.

### Building the simulated world

The simulated world consists of different types of locations where agents can stay and between which agents move. The types of locations implemented in the model are agents’ homes, workplaces, school classes, kindergartens, universities and supermarkets, in order to simulate the most important NPIs that had come into effect in each federal state in Germany in Spring 2020. The subsequent NPIs have been closed schools, kindergartens and universities, increased usage of working from home (yet not at full scale), quarantine of the household of infected people and reduced working hours due to the economic shutdown.

For each household of the population a home is created in which all members of the household (taken from the SOEP) reside. The number of additional location types is adjusted to the characteristics of the agent population and the respective federal state. Table 1 (cf. SI) shows the parameters we use to calculate the corresponding number of locations in the simulation model.

### Daily routines

During the day, agents engage in various activities. An activity is implemented in the simulation as staying at a certain place for a certain period of time. The type of activity an agent engages in, when and for how long, depends on the clock time, the weekday, the agent’s attributes and the NPIs that are currently in effect. The default activity that every agent exhibits when there is no other activity on its schedule is being at home. The simulation starts with every agent being at home. Each time step represents an hour. Every morning at 8 o’clock each agent first decides whether it should stay at home due to symptoms of an infection or household quarantine. If neither is the case, if the agent’s daily schedule is not affected by an NPI and if it is a weekday from Monday to Friday, kindergarten kids go to kindergarten, school kids go to school, university students attend university, and workers go to work. To account for the possibility of home office in the simulation, working agents change their location to their workplace only with a certain probability and otherwise start working from home. The probabilities to work from home are derived from data on the frequency of home office use per NACE-category in Germany in 2018 [4]. The respective NPI on increased home office usage sets numbers as if the home office capacity would have been exploited in each NACE-industry [4]. Therefore, the scenario in which home office is reduced to pre-pandemic times must be interpreted accordingly.

School kids and kindergarten kids return to their homes after 5 hours, university students after 4. If university students have jobs, they first go to work and after work they go to university. While the dwell time at kindergarten, school and university are based on assumptions, the daily working hours represent those of the modeled survey respondents. When agents are at home and do no other activity, there is a certain probability to go to the supermarket. Every night at 1 a.m. the clock is set forward to 7 a.m. so that most of the night time is skipped.

### Modeling the infection

The biological aspect of the infection, i.e. what happens to an agent after the virus was successfully transmitted, was implemented in strong orientation to the way the process of infection is implemented in “Covasim” [28]. It can be considered as an extended S-E-I-R (susceptible, exposed, infectious, recovered) infection model [2]. If an infectious agent and a susceptible agent are at the same time in the same place, there is certain probability of infecting the susceptible agent. At each time step, each infectious agent randomly selects one agent that is currently in the same location if the infectious agent is currently not alone in its location. If the agent randomly selected by the infectious agent is susceptible to infection, its infection status is set to ‘exposed’ by a certain probability of infection. This hourly probability of infection is a very important, but unknown parameter and is therefore determined by model calibration to empirical infection data of a given state (cf. section “Scaling and calibration”). How long each stage lasts and if the agent develops symptoms or not, is determined individually for each agent based on its empirical characteristics. For each agent and each stage of infection we draw individual time spans from a log-normal-distribution following Covasim [28]. In addition, the decision whether an agent develops symptoms or not is determined by an age-dependent probability, which also resembles Covasim [28].

### NPIs and scenarios

The initial goal of the model is to capture the course of the pandemic during its first wave in Germany. For each state, we simulate 100 days, starting with the day when a state had approximately 50 cases per 100,000 inhabitants. While this defines the scope of the paper’s results, in future research the model could also be extended to further waves to come. The main aim is to estimate effects of multiple NPIs implemented during the first wave in Germany. These effects must be considered in the model in order to capture the mechanisms of virus spread as close to reality as possible. The changes on the agents’ daily lives due to NPIs considered in the model are the following: less to no time at school, kindergarten or university, an increased amount of time working from home, quarantining households of infected agents and reduced or no work hours due a (partial) shutdown of certain branches of industry.

Most effects of NPIs are implemented as a change in probability of actually leaving the home and exercising a certain activity. In response to active NPIs, the specific probability reflects whether a certain location is closed (or to what degree), or that work hours are reduced for a given NACE-category. For example, a (partially) closed school is implemented as a daily probability that determines for each school kid separately whether he or she goes to school that day. If schools are completely open, the probability for school attendance is 1. Complete closure of schools are reflected in a daily probability of school attendance equal to 0. If an agent shows symptoms of an infection over a certain period of time, the agent and all household members are quarantined at home for 14 days.

In the baseline model we mimic the course of the lockdown in Germany with its onset at around march 15 and the following 3 months of disease spread. It is important to note that each effect is the change in the number of cases if a given NPI had not been implemented while all other NPIs remain active (if not reported differently, e.g., the combined effects of education-related NPIs in the “School Kinder”-scenario).

### Scaling and calibration

In each run of the simulation we collect the new infections per day, the cumulative cases on a day and the age of the infected agents. If the size of the agent population is exactly equal to 100000 cases, frequencies can be directly interpreted as cases per 100000 inhabitants. Otherwise, the infection frequencies are calculated by *I ∗*100000*/P*, where I is the number of infections and P is the actual population size in the simulation. Therefore, all results can be interpreted as cases per 100000.

Due to the many stochastic elements used in the simulation, each single run of the model returns different results, even when the same set of parameters is used. Thus, to obtain reliable results for a given set of parameters we run 60 repetitions of model runs and average the results.

Although the model includes relevant micro-level mechanisms of a pandemic and a representative agent population, we must perform some iterative fine-tuning as is often done in ABMs, i.e., calibrating the model to reproduce empirical data [45]. To calibrate the ABMs on infection frequencies in a given federal state, we determine the values of two parameters: the hourly probability an infected agent will infect another agent, and the time needed to quarantine a household after the first symptoms of a household member appear. We performed the calibration using the “Latin Hypercube Sampling” algorithm [45], which is implemented in the python package “Spotpy” [25]. For each model to be calibrated, we searched for the set of model parameters that generate simulated data with the smallest deviation to the corresponding daily empirical data on cumulative number of infections (data source: [42]). We stopped the calibration process when at least one set of parameters was found to generate simulated data with a fit of a root mean squared error less than 20. Because the calibration of the model for a single federal state requires at least 180 simulation runs (each consisting itself of 60 internal runs and one run takes approx. 20 minutes) due to varying parameter sets, each process of calibration was serialized on 192 cores provided by BW-HPC, a statewide computing and data infrastructure.

## Data Availability

The empirical calibration of our model is largely based on survey data from the Socio-Economic Panel. Unfortunately, we are not allowed to share this dataset due to a highly restrictive data access policy. However, as a member of an academic institution, you can request access to the datasets from DIW Berlin (https://www.diw.de/soep). All other datasets used in this study are freely available to the public. The complete Python code and all STATA-Do-Files will be published on GitHub.

## A Supplementary information

**Table 1:** Values and data sources of simulation parameters

| Parameter | Value | Source |
| --- | --- | --- |
| Hours worked on weekdays | agent-specific | [43] |
| Age | agent-specific | [43] |
| Gender | agent-specific | [43] |
| Federal state | agent-specific | [43] |
| Nace rev. 2 category | agent-specific | [43] |
| Household weight | household-specific | [43] |
| Enrollment at the university | agent-specific | [43] |
| Daily hours at stores | agent-specific | [43] |
| Daily hours at kindergarten | 5 | Assumption |
| Daily hours at school | 5 | Assumption |
| Daily hours at universities | 4 | Assumption |
| Kindergarten age | 0 - 5 | Assumption |
| School age | 6 - 19 | Assumption |
| Supermarkets per agents | 1/1000 | Assumption |
| Colleagues per work place | 10 | Assumption |
| Number of supermarkets an agent visits | 2 | Assumption |
| Number of Universities | 1 | Assumption |
| Days until an agent cures symptoms at home | 1 | Assumption |
| Duration of household quarantine in days | 14 | Assumption |
| Daytime | 8 - 21 | Assumption |
| Probability of developing symptoms at age 0-9 | 0.5 | [28] |
| Probability of developing symptoms at age 10-19 | 0.55 | [28] |
| Probability of developing symptoms at age 20-29 | 0.6 | [28] |
| Probability of developing symptoms at age 30-39 | 0.65 | [28] |
| Probability of developing symptoms at age 40-49 | 0.7 | [28] |
| Probability of developing symptoms at age 50-59 | 0.75 | [28] |
| Probability of developing symptoms at age 60-69 | 0.8 | [28] |
| Probability of developing symptoms at age 70-79 | 0.85 | [28] |
| Probability of developing symptoms at age > 80 | 0.9 | [28] |
| Days from exposed to presymp. infectious | lognormal(4.6, 4.8) | [28] |
| Days from presymp. infectious to (a)symp. infectious | lognormal(1, 1) | [28] |
| Days from (a)symptomatic to recovered | lognormal(8, 2) | [28] |
| Average class size Baden-Wuerttemberg | 22 | [15] |
| Average class size Bavaria | 23 | [15] |
| Average class size Hamburg | 25 | [15] |
| Average class size Saarland | 24 | [15] |
| Average kindergarten group size in Baden-Wuerttemberg | 15 | [5] |
| Average kindergarten group size in Bavaria | 18 | [5] |
| Average kindergarten group size in Hamburg | 18 | [5] |
| Average kindergarten group size in Saarland | 18 | [5] |

**Table 2:** Calibrated parameters

| Parameter | Baden-Wuerttemberg | Bavaria | Hamburg | Saarland |
| --- | --- | --- | --- | --- |
| Hourly infection probability | 0.06213 | 0.0645 | 0.0502 | 0.04727 |
| Days from symptoms to household quarantine | 1.21 | 1.32 | 1.40 | 1.30 |

**Table 3:** Descriptive statistics on federal states

| Variable | Baden-Wuertt. | Bavaria | Hamburg | Saarland | Source |
| --- | --- | --- | --- | --- | --- |
| GDP 2019 in billions € | 524.33 | 632.90 | 123.27 | 36.25 | [1] |
| Inhabitants per km<sup>2</sup> 2018 | 310 | 185 | 2438 | 385 | [16] |
| Average age in agent pop. | 46.153958 | 45.791286 | 45.164004 | 44.727600 | [43] |
| Average household size in agent pop. | 2.791733 | 2.605203 | 2.735912 | 2.513871 | [43] |
| Average work hours in agent pop. | 3.091075 | 3.106836 | 2.696887 | 3.082421 | [43] |
| Most frequent NACE Rev.2 in agent pop. | C (14.62%) | C (13.02%) | Q (7.94%) | Q (12.72%) | [43] |
| Men in agent pop. | 48.33% | 48.47% | 44.86% | 46.44% | [43] |
| Pupils in agent pop. | 13.31% | 12.53% | 14.44% | 12.65% | [43] |
| Kindergarten kids in agent pop. | 5.01% | 4.81% | 5.08% | 4.51% | [43] |
| University students in agent pop. | 2.32% | 2.12% | 3.44% | 1.45% | [43] |

**Figure 5:**
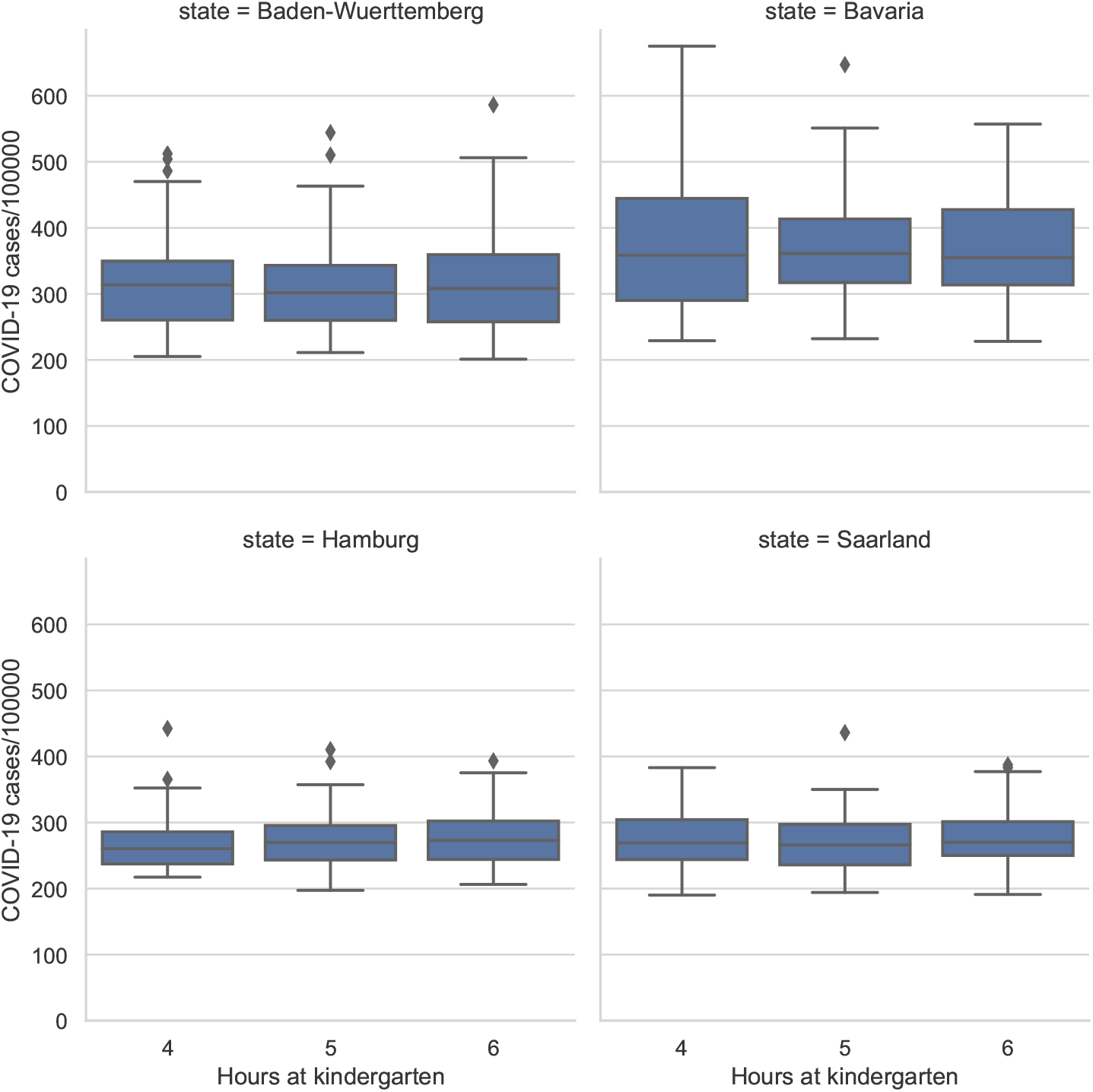
Robustness check: hours at kindergarten. Compares the number of COVID-19 infections in the baseline scenario across different values for the daily hours at kindergarten. The value in the center is the one used in the main analysis.

**Figure 6:**
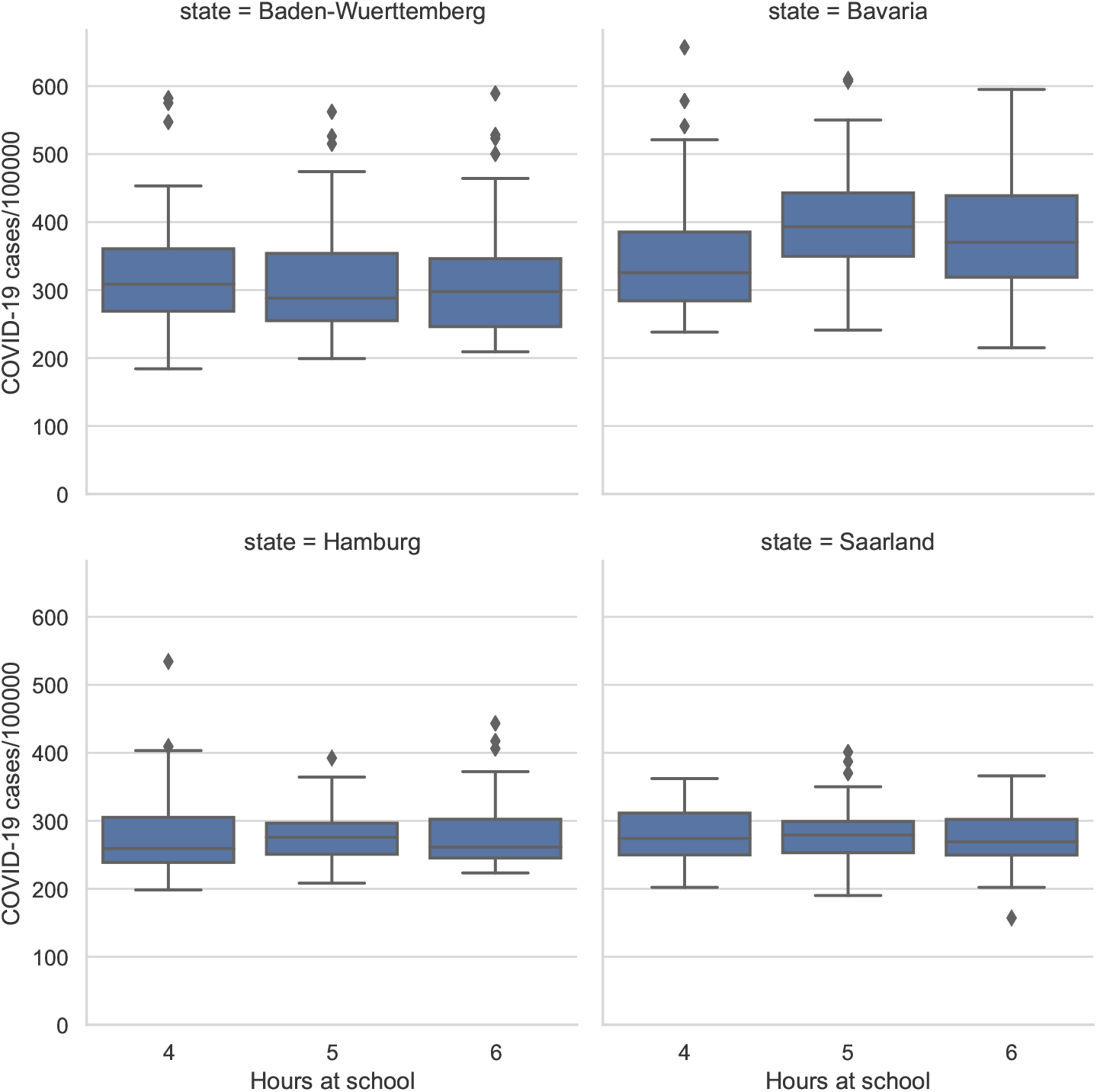
Robustness check: hours at school. Compares the number of COVID-19 infections in the baseline scenario across different values for hours at school. The value in the center is the one used in the main analysis.

**Figure 7:**
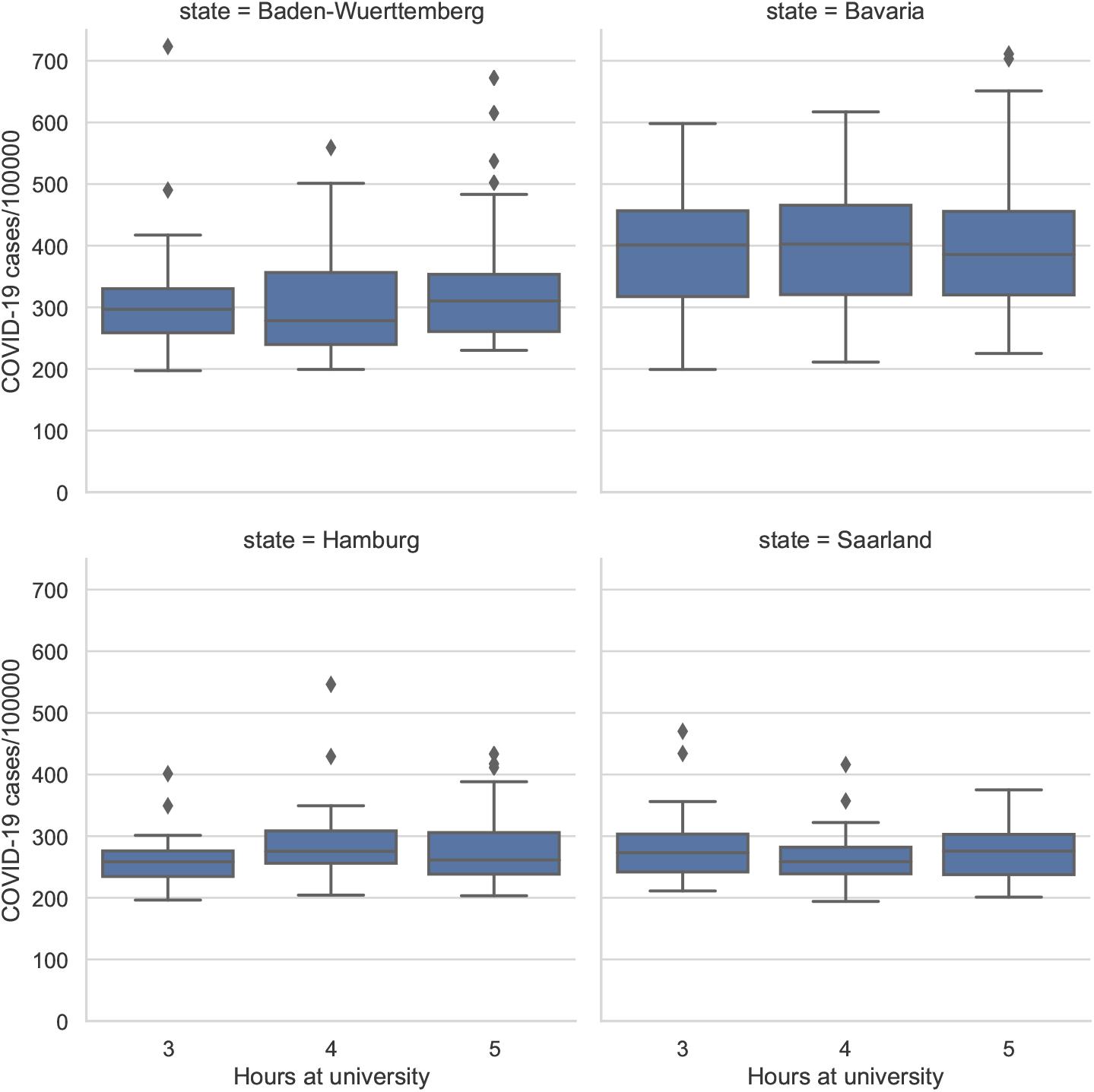
Robustness check: hours at university. Compares the number of COVID-19 infections in the baseline scenario across different values for hours at university. The value in the center is the one used in the main analysis.

**Figure 8:**
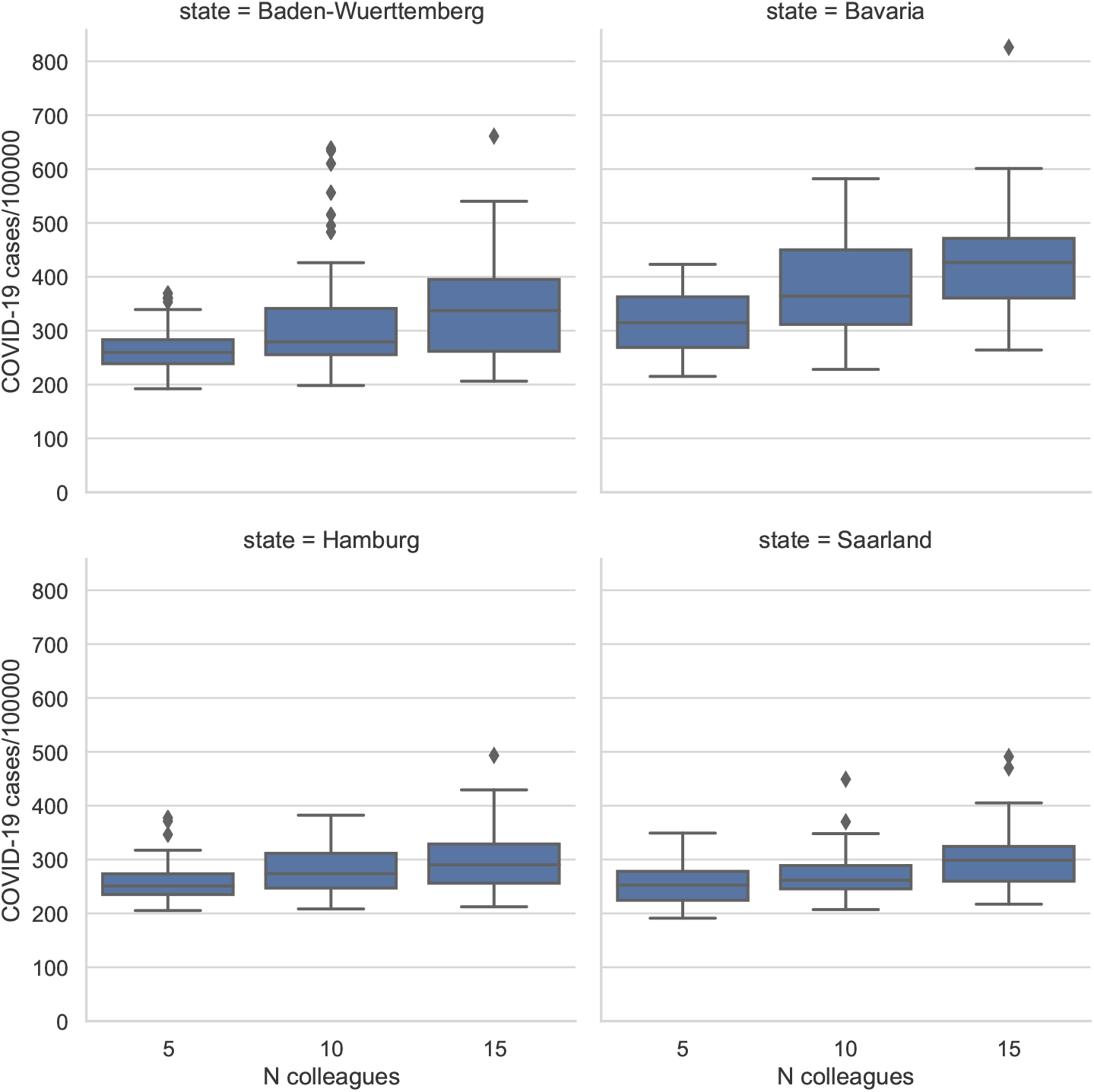
Robustness check: number of colleagues. Compares the number of COVID-19 infections in the baseline scenario across different values for number of colleagues. The value in the center is the one used in the main analysis.

**Figure 9:**
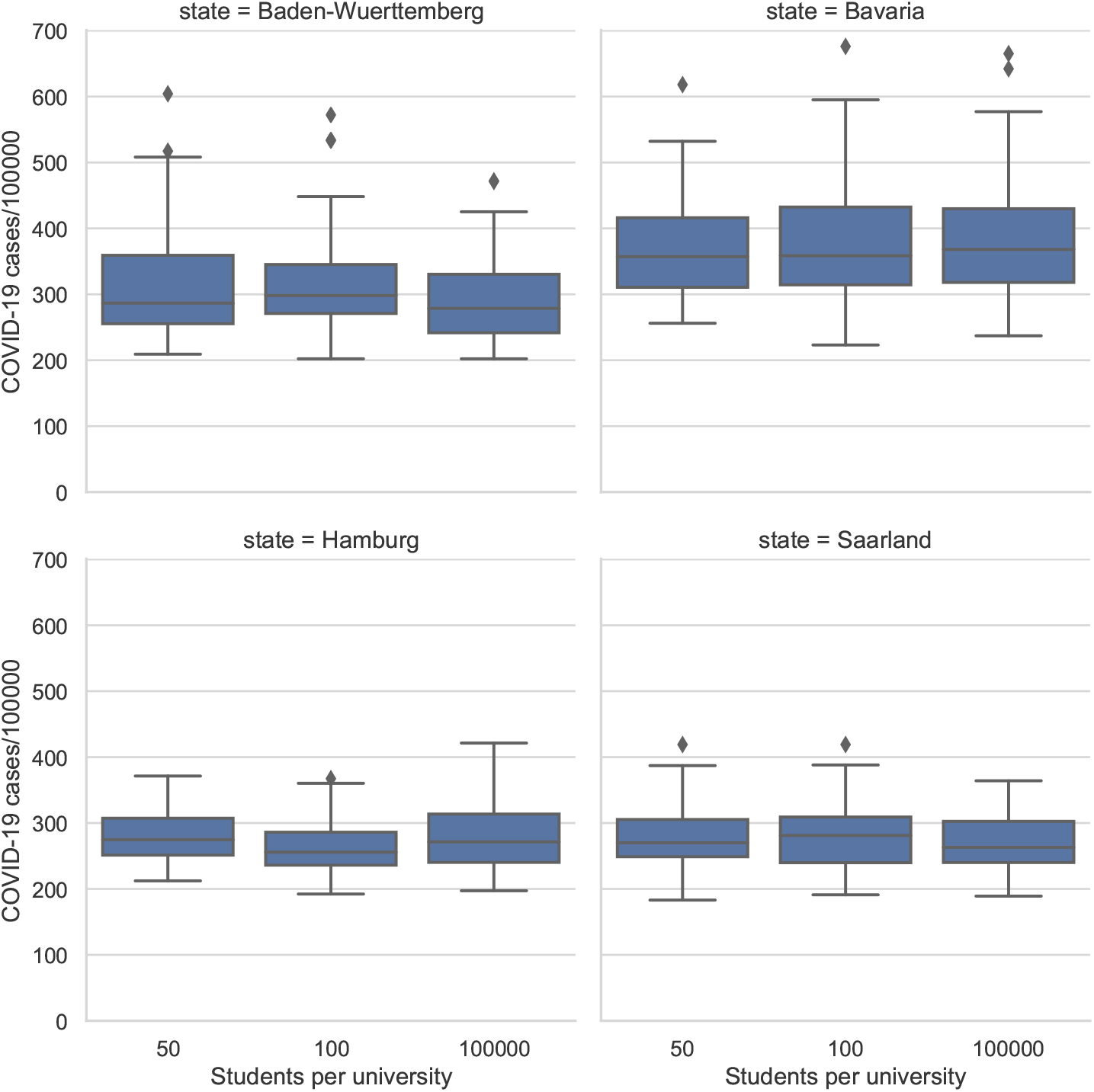
Robustness check: maximum number of students per university. Compares the number of COVID-19 infections in the baseline scenario across different values for the maximum number of students per university. We used 100,000 in the main analysis, which means that there is only one university per simulated world.

## Footnotes

1 All code is made available to reproduce results or to add further scenarios and, respectively, NPIs.

